# Performance of automated anterior segment OCT-based quantitative imaging in adult anterior chamber inflammation

**DOI:** 10.64898/2026.07.06.26357402

**Authors:** Ameenat L Solebo, Boyu Chen, Nik Khadijah Nik Aznan, Miguel Xochicale, Tom Roberts, Harry Petrushkin, Claire Lim, Rongling Shu, Megan Jacobson, Idil Farisogullari, Giuseppe Demarinis, Khaled Abdelfattah, Dominique Tynan, Jivan Lotay, Kanwaldeep Vijan, Chrysanthi Tsika, Olayinka Williams, Gerry Clare, Ilaria Testi, William R Tucker, Peter Addison, Edmund Tsui, Carlos Pavesio, Jugnoo Rahi, Paul Taylor, Colin J Chu

## Abstract

**Objective:** To investigate the performance of anterior segment (AS-) OCT quantitative imaging of anterior chamber inflammation in uveitis patients with diverse demographics.

**Design:** Prospective cross-sectional study.

**Participants:** 144 adult patients managed at a tertiary care service in the UK

**Methods:** Repeated swept-source AS-OCT imaging was performed pre- and post-pupil dilation (i.e. 4 scan sets). Inflammation was quantified using a validated human-in-the-loop automated image analysis pipeline, Minuscule Cell Detection (MCD), which identified and counted putative inflammatory cells on AS-OCT.

**Main Outcome Measures:** Test-retest variability of AS-OCT and diagnostic accuracy of various AS-OCT derived measurands (minimum, maximum, median counts per cross sectional image, and total counts across volume image sets per eye or MINCC, MAXCC, MEDCC and TOTCC) versus Standardization of Uveitis Nomenclature (SUN) grading system as assessed by a uveitis specialist.

**Results:** A total of 281 eyes were included in the analysis. Median age was 48 years (IQR 36-64). Strong test-retest measurand reliability was demonstrated, with a 95% tolerance interval ratio 0.3 - 3.0. The best diagnostic performances for SUN activity were observed with the MINCC threshold of 3 particles (negative predictive value for clinical activity of 89.8%, 95% CI 83.0 - 94.1). Associations between AS-OCT measurands and patient age (adjusted coefficient 7.5 additional particles, 95% CI 0.5 – 14.6, p<0.04 for age over 60 years versus under 44), and pigment load (52.8, 11.8 - 92.9, p<0.01 in eyes with AC pigment versus without) were noted.

**Conclusions:** AS-OCT assessment of anterior chamber inflammation in uveitis meets current recommendations for quantitative imaging biomarkers, demonstrating strong repeatability, linearity with clinical assessment scores and stability with pupil dilation and patient characteristics of ethnicity and lens status. The absence of variability in diagnostic indices across derived measurands suggests similar performance across different acquisition protocols. Further longitudinal cross-platform studies are needed to determine limitations of use.

---

Uveitis is a globally significant cause of vision loss in working age adulthood,^1^ and frequently involves the ocular anterior segment. In addition to its impact on health outcomes, uveitis is a significant driver of health care service utilization: anterior uveitis is one of the most common reasons for attending an acute eye care centre.^2^ The diagnosis and ongoing monitoring of the chronic and recurrent forms of disease also impose a considerable burden on health care systems due to repeated visits for specialized care.^3^ Timely, sight-saving care of this disorder is threatened by the global, worsening shortage of ophthalmic professionals^4,5^

Currently, anterior uveitis is diagnosed following slit lamp examination by a suitably trained professional.^6^ Active disease may be missed by less experienced clinicians.^7,8^ Anterior segment optical coherence tomography (AS-OCT) has the potential to be an objective, sensitive disease tool for diagnosis and monitoring across hospital and community settings. Previous investigators have focused on the clinical validation of quantitative AS-OCT imaging for intraocular inflammation, measuring the correlation of AS-OCT findings with clinical assessment,^9–11^ but we lack the robust analytic validation necessary for clinical adoption.^12,13^ There is little evidence on whether AS-OCT, as a quantitative imaging technology for assessing anterior chamber inflammation, produces consistent results, or how performance varies across conditions and populations.

We evaluated the characteristics of AS-OCT quantification of anterior segment inflammation in adults with a mixed real-world cohort, including repeatability,^13^ limits of quantitation, and the impact of patient characteristics such as age, sex, ethnicity and co-morbidity (lens status and anterior chamber pigment).

## METHODS

This cross-sectional study prospectively recruited adults (aged >18 years) managed at a single tertiary care Uveitis service (Moorfields Eye Hospital). Individuals with corneal opacity sufficient to obscure slit lamp grading of inflammation in the anterior chamber were excluded. Patients attending clinics were directly approached for recruitment between October 2024 and February 2025.

### Image acquisition for index testing

Images of both eyes of each participant were acquired pre and post pupil dilation using the swept source AS-OCT, Anterion model (Heidelberg Engineering, Heidelberg, Germany, 1300-nm light source and scanning speed of 50,000 A-scans/second), using a raster centered at the pupil center. Images were acquired by 8 operators at one operating site, following confirmation of correct operating technique given by one of the two senior authors (ALS, CC). Full acquisition settings are available in supplementary document SD1, but, in brief, comprised a 14mm scan length, 6mm height, 9-line horizontal B-scan raster with 768 A-scans per B-scan and automated real-time (ART) of 1. Each eye of every individual was imaged at least four times: twice prior to dilation (PreD1 and PreD2), with a 30-60 second interval, and twice post-dilation (PostD1 and PostD2, again separated by 30-60 seconds). Images from all line scans were extracted and converted into PNG format for analysis.

### Image analysis

The resulting PNG images enabled a standardized downstream analysis by applying an automated cell quantification approach with the Minuscule Cell Detection (MCD) pipeline (code available at GitHub - joeybyc/MCD),^14^ which was used to identify and count putative inflammatory cells within the anterior chamber. Prior to running inference across the full dataset, an image-level analysis was performed to optimize inference parameters for the images. Full details of this process are available in supplementary document SD2. Inference was then run across the entire dataset with detected cells counted on a per-B-scan basis. For each B-scan, MCD first leverages a zero-shot vision foundation model to segment the AC region. Within this region, MCD then employs a Minuscule Region Proposal (MiRP) algorithm to identify candidate regions likely to contain cells. These regions are cropped as small, cell-sized patches for the subsequent stage, avoiding the need to process the full image. Each candidate patch is then passed through a lightweight neural network, which learns discriminative local features to distinguish true inflammatory cells from background speckle noise and imaging artefacts.

Detected cells were subsequently counted on a per-B-scan basis (figure 1). To derive a robust eye-level estimate, analysis was restricted to B-scans 3, 4, and 6 (out of nine available slices per eye, with the central scan, 5, excluded due to the light glare which is the result of the fixation beam), which consistently captured the central anterior chamber region (figure 2). This resulted in a clear monotonic relationship between the manually annotated and predicted cell counts (supplementary figure SF1).

**Figure 1.**
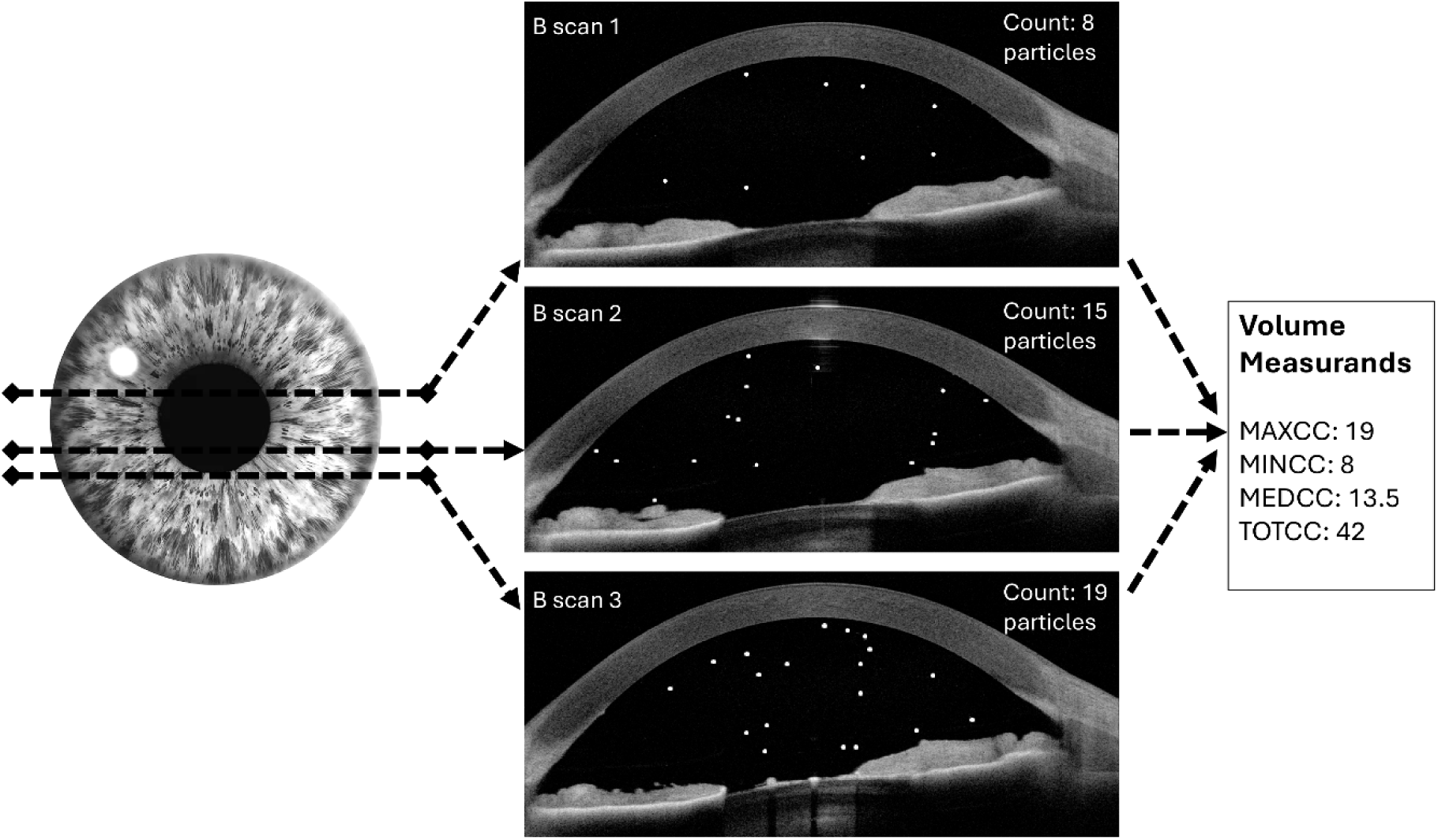
Comparison of clinical scores and automated particle count of clinically active and inactive examples. MCD=automated anterior chamber particle quantification using the Minuscule Cell Detection pipeline SUN=Standardized Uveitis Nomenclature anterior chamber cell score

**Figure 2.**
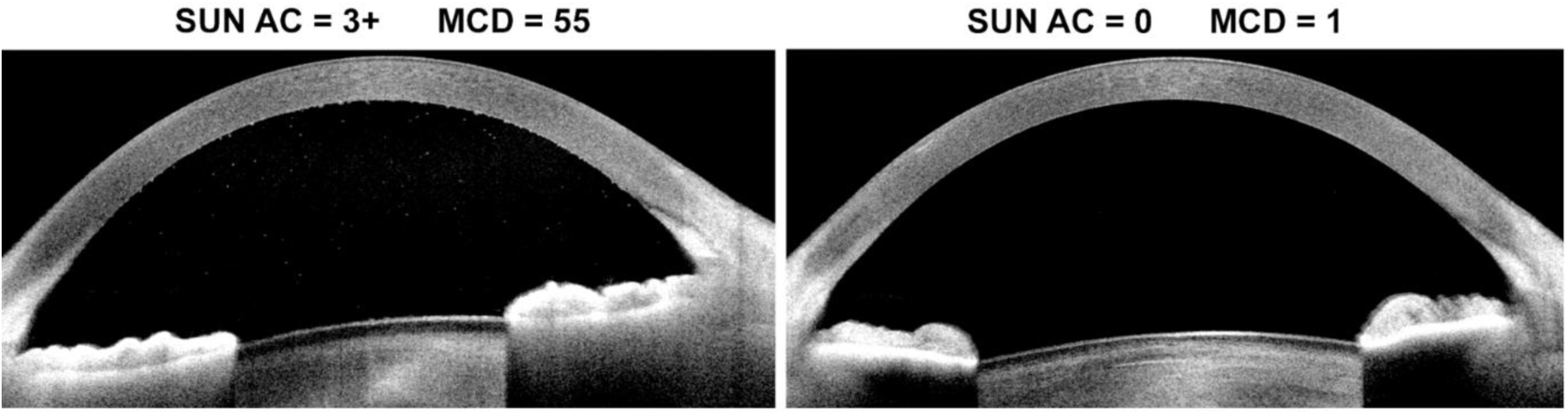
Schematic of measurand generation. The central most B-scan was excluded from analyses due to the fixation light glare artefact present in all central scans

### Measurand generation

Since future routine use of AS-OCT might be limited to the acquisition of a single cross-sectional scan, we generated multiple potential measures (termed measurands, for consistency with the US Food and Drug Administration recommendations on quantitative metrics in health)^13^ from each volume. This enabled a deeper understanding of the expected variability within as well as across volumes. In recognition of the variety of future use cases for this technology in clinical care, the following imaging-based measurands (i.e., properties measured in each scan) were derived from the central three scans from each volume: MAXCC, or maximum cell count per horizontal cross-sectional scan per volume, in order to capture the ‘most active’ scan acquired from each eye; MINCC, or minimum cell count per cross-sectional scan, in order to capture the ‘least active’ scan, and the scan most likely to be most easily and precisely assessed by a human observer in future clinical adoption;^15^ TOTCC, or total cell count across all three central scans, i.e. a summary score which used the maximum data available, but would be dependent on the number of scans analyzed; and MEDCC, or median cell count per cross-sectional scan, i.e. a summary quantification score more easily extrapolatable to future volume sets across different acquisition protocols (figure 2).

### Analytic validation of index testing

To generate otherwise unavailable information on the limits of precision for AS-OCT capture of inflammation severity, test-retest variability was assessed using Bland-Altman analyses of the measurands generated from the PreD1 versus the PreD2 scans, and again, separately, from the PostD1 versus PostD2 scans. The impact of dilation on the measurands was tested using Bland-Altman analyses of the PreD1 and PostD1 scans, with multivariable regression analyses to further interrogate the impact of age, sex, race / ethnicity, lens status, activity status and anterior chamber pigment load on repeatability of AS-OCT quantification pre and post dilation.

### Clinical reference (clinical bio-microscopy) test

Prior to pupil dilation, and prior to the first set of imaging, patients underwent examination by a uveitis sub-specialist fellow or a uveitis nurse practitioner, who also examined the eye post dilation (again prior to the post dilation AS-OCT). Pre-dilation examination was also undertaken by a consultant (attending) uveitis specialist. The attending clinicians were masked to the findings of the other clinician, and all clinicians were masked to index test findings. Anterior chamber (AC) inflammation was assessed using the ordinal standardization of uveitis nomenclature (SUN) grading system.(10) Clinically active disease was defined as a SUN grading of ≥+1, or at least 6 cells seen in a 1 mm × 1 mm beam. SUN grading was also used to assess AC flare, and a three-level grade was used to capture the level of AC pigment dispersion (none, moderate, heavy).^11^ To quantify the precision of clinical assessment (as the ‘gold standard’ test for diagnostic accuracy) agreement between clinicians was assessed using intraclass correlation coefficient analyses, with kappa scores weighted to penalize 2 SUN grade steps or larger disagreements in clinical grading.

### Clinical validation

Correlations between clinical SUN score for anterior chamber cell activity (reference test) and measurand (index test) were assessed using logistic regression (with SUN score as the dependent variable). The impact of participant characteristics (sex, age, race / ethnicity and lens status) on the relationship between reference and index testing were assessed using multivariable regression models, with multilevel adjustment for the within-individual correlation of eye-level data. Correlations between potential covariables were investigated using nonparametric tests (χ2, Mann–Whitney U, and Spearman), with a *p* value threshold of 0.05 selected as indicative of a statistically significant correlation. Models were constructed using conventional forward and backward stepwise regression and included variables significant at a 10% level in initial univariable analysis. Where there was correlation between independent variables (*p*<0.05), relationships were explored using interaction terms. Where only one variable was found to reach statistical significance (*p*<0.10) on univariable modelling, adjustment was made for the other putative correlated factors. Factors were retained in the multivariable model if they altered the estimate by more than 10%, were independently associated at a 5% significance level, or if there was no significant correlation between covariables.

Conventional two by two tables were used to evaluate the diagnostic accuracy (including sensitivity, specificity and likelihood ratios) of selected measurands thresholds. A clinically positive diagnosis was defined as ≥SUN +1 for anterior chamber cells as assessed by the consultant / attending pre-dilation. The measurand thresholds were selected using exploration of the 1^st^, 5^th^, and 10^th^ centiles for each measurand from images of eyes determined clinically to be active at the slit lamp. Receiver operating characteristic curves (using data from all eyes, with correction for data clustered within the individuals) were then plotted to explore the diagnostic power of each measurand.

Statistical analyses were undertaken in Stata version 19.5 (StataCorp).

### Informed Consent

Informed and written consent was obtained from all participants.

### IRB/Ethics Committee

Ethics Committee approval was obtained from the United Kingdom Health Research Authority (South Central Oxford B Research Ethics Committee, reference 19/SC/0283). This study adhered to the tenets of the Declaration of Helsinki.

## RESULTS

AS-OCT images were acquired from 156 adults with uveitis. Of the 156 adults, seven participants had only one eye eligible for inclusion. A complete set of sufficient quality images were achieved from 281 (139 right eyes, 142 left eyes, 144 individuals) of the 305 eligible eyes. In total, 2,529 B-scans were processed by the human-in-the-loop automated MCD pipeline. Median age of included participants was 48 years, (range 22 to 84, interquartile range, IQR, 36 to 64 years). Patient characteristics for eyes included in analyses are presented in Table 1. ^14^

**Table 1.** Characteristics of included eyes and participants.

|  |  | Total<br>n=281 eyes (%) [144<br>participants] |
| --- | --- | --- |
| Gender: |  |  |
|  | Female | 169 (60.6%) |
|  | Male | 110 (39.1%) |
|  | Missing | 3 [2 participants] |
| Self-reported ethnicity: |  |  |
|  | White British / Other | 137 (48.8%) |
|  | Asian / Asian British | 38 (13.5%) |
|  | Black / Black British: | 71 (25.3%) |
|  | Other | 29 (10.3%) |
|  | Not reported | 6 [3 participants] |
| Age: |  |  |
|  | <44 years | 105 (37.4%) |
|  | 44 – 60 years | 82 (29.2%) |
|  | >60 years | 94 (33.5%) |
|  | Missing | 0 |
| Uveitis type: |  |  |
|  | Anterior | 150 (53.4%) |
|  | Intermediate | 33 (11.7%) |
|  | Panuveitis | 50 (17.8%) |
|  | Posterior | 48 (17.1%) |
|  | Missing | 0 |
| Lens status: |  |  |
|  | Phakic | 183 (65.1%) |
|  | PC-IOL | 91 (32.4%) |
|  | AC-IOL | 5 (1.8%) |
|  | Aphakic | 2 (<1%) |
|  | Missing | 0 |
| Anterior chamber cell grade (SUN)*: |  |  |
|  | 0 | 197 (70.1%) |
|  | +0.5 | 34 (12.1%) |
|  | +1 | 21 (7.5%) |
|  | +2 | 23 (8.2%) |
|  | +3 or more | 6 (2.1%) |
| Anterior chamber flare grade (SUN)*: |  |  |
|  | 0 | 196 (69.8%) |
|  | 1 | 77 (27.4%) |
|  | 2 or more | 8 (2.8%) |
| Anterior chamber pigment density*: |  |  |
|  | None | 246 (87.5%) |
|  | Moderate | 32 (11.4%) |
|  | Heavy | 3 (1.1%) |

AC: Anterior chamber; PC: Posterior chamber; SUN: Standardization of Uveitis
\*as assessed pre-dilation

### Measurand scores

MAXCC, or maximum cell count per horizontal cross-sectional scan per volume scores ranged from 1 - 547, median 5; MINCC, or minimum cell count per cross-sectional scan per volume ranged from 0-227, median 2; MEDCC, or median cell count per cross-sectional scan across a volume ranged from 0-482, median 5; and TOTCC, or total cell count across scans ranged from 1-1256, median 10.

### Test-retest variability of AS-OCT for cell counts

A critical factor to be determined for wider clinical deployment, there was strong reliability of generated counts on Bland-Altman analysis of sequential scans (same eye, prior to dilation, with scans separated by 30-60 seconds) (Figure 3). Overall, the 95% confidence interval of MAXCC counts ran from -14.7 (the first scan having approximately 15 particles fewer in the most active slice) to 13.7 (the first scan having 14 particles more). These data are presented graphically as Bland-Altman plots in supplementary figure SF2. Following correction for the number of particles within scans, the 95% tolerance interval ratio with 95% confidence (95% TIC) for MAXCC (i.e. the ratio range within which 95% of future individual differences are expected to fall with 95% confidence) was 0.3 to 3.0. This three-fold difference in the 95% TIC as a limit of variability, outside which a change in particle count would be more likely to represent a ‘true change’, remained robust across the different measurands: MINCC 0.3 to 3.9 (uncorrected 95% CI for test-retest -9.0 to 9.7 particles); MEDCC 0.4 to 2.6 (-25.1 to 23.3); and for TOTCC, 0.3 to 3.2 (-26.3 to 26.1). Sequential post-dilation scans also demonstrated a three-fold difference between counts as an expected limit for the stability of future tests, with a 95% TIC for MAXCC of 0.3 to 3.2; MINCC 0.3 to 3.0; MEDCC 0.4 to 2.7; and for TOTCC, 0.3 to 3.3.

**Figure 3.**
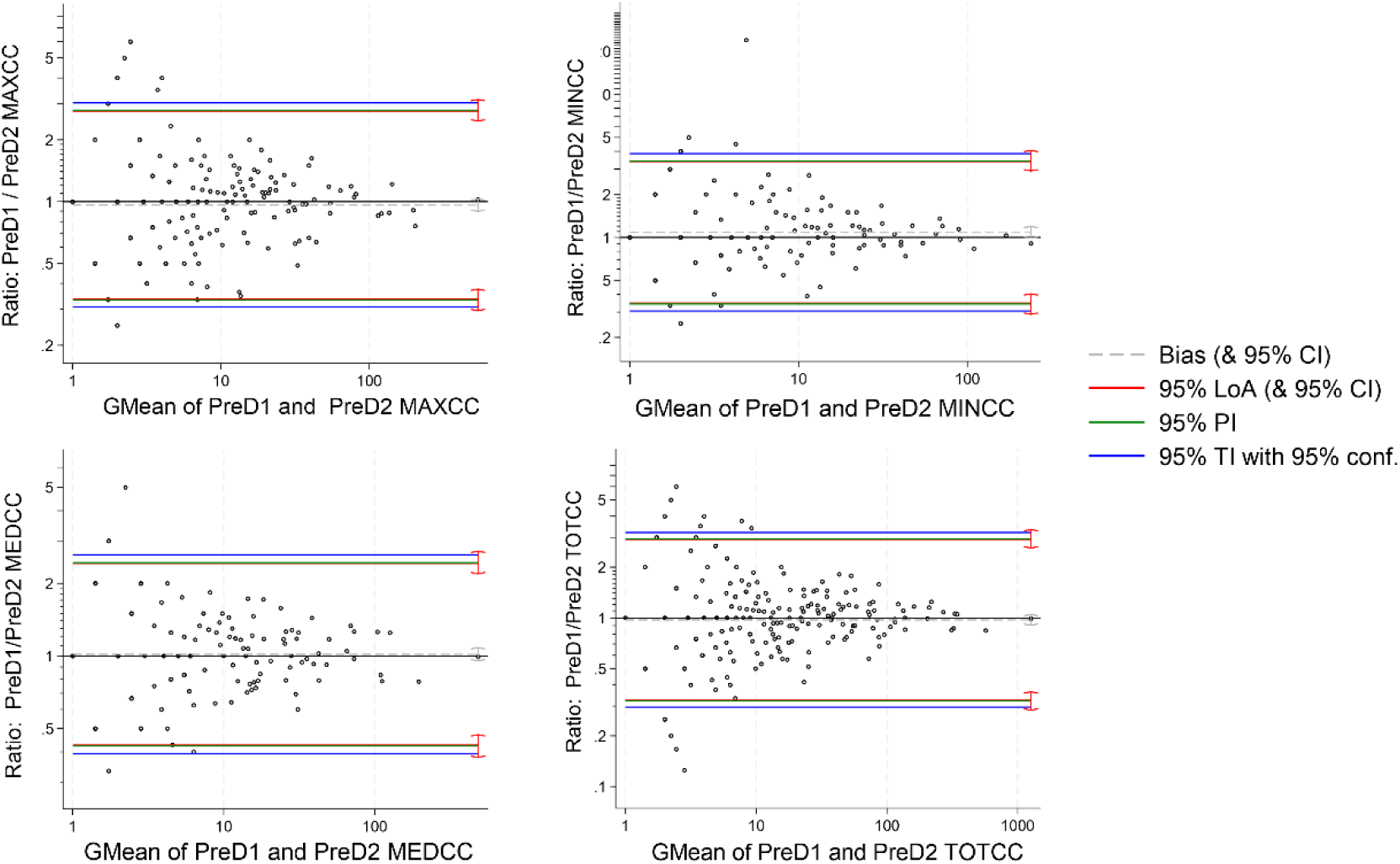
Bland-Altman plots for test-retest variability of measurands. MAXCC: maximum cell count per horizontal cross-sectional scan per volume; MINCC: minimum cell count per cross-sectional scan; MEDCC: median cell count per cross-sectional scan; TOTCC: total cell count per volume; GMean: geometric mean; LoA: limits of analysis; PI: performance interval; TI: tolerance interval Measurand geometric means and ratios have been used to account for the non-parametric (right-skewed) structure of the count data.

### Impact of pupil dilation on AS-OCT cell counts

On comparison of pre versus post dilation, there was a tendency for post dilation counts to be higher across all measurands (presented as scatter plot data in supplementary figure SF3) but this did not reach statistical significance, and Bland-Altman analyses did not demonstrate a significant bias to test-retest variability.

### Clinical assessment of AC activity demonstrates high intergrader agreement

Prior to analyses of diagnostic accuracy, evaluation of slit lamp clinical assessment metrics provided confidence in the ‘gold standard’ to be used as a comparator against AS-OCT counts. Agreement between clinicians (consultant / attending versus residents) was substantial for slit lamp anterior chamber (AC) cells SUN grading (weighted kappa of 0.62), and fair for AC pigment density (0.31). Consultant / attending assessments were used consistently as the gold standard for diagnostic accuracy.

### Validation of AS-OCT scores against clinical assessment

There was a robust difference in all AS-OCT measurand counts for inactive versus active uveitis eyes. Multilevel receiver operating characteristic curves analyses were undertaken to compare the diagnostic performance of the different potential measurand thresholds. This enabled the selection of the best performing thresholds of: MAXCC=6 (disease activity indicated by 6 or more particles in any cross-sectional scan), MINCC=3 (activity indicated by no fewer than 3 particles in any cross-sectional scan) and MEDCC=5 (activity indicated by median of 5 or more particles per cross sectional scan).

Two by two tables (supplementary table ST1) identified strong diagnostic performance irrespective of which measurand threshold was selected, with successful identification of all cases of SUN 2+ or greater levels of activity. The best negative predictive values (NPV, best at ‘ruling out’ disease activity) were achieved with MINCC=3, NPV 89.84% (95% CI 83.01% to 94.12%) whilst the lowest false positive error rates were seen with MEDCC=5, positive predictive value 63.42% (95% CI 51.78% to 73.68%) (table 2). Receiver operated curve characteristics, demonstrating the strong diagnostic performance across the selected measurand thresholds, are presented in supplementary figures SF4.

**Table 2.** Diagnostic performance of selected measurand thresholds for clinical activity.

|  | MINCC=3 | MAXCC=6 | MEDCC=5 |
| --- | --- | --- | --- |
| Accuracy | 78.13% | 76.56% | 79.23% |
| 95% CI | 70.14% to 84.83% | 68.43% to 83.47% | 71.34% to 85.78% |
| Sensitivity | 79.55% | 79.55% | 72.73% |
| 95% CI | 64.70% to 90.20% | 64.70% to 90.20% | 57.21% to 85.04% |
| Specificity | 77.53% | 75.28% | 82.02% |
| 95% CI | 67.45% to 85.70% | 65.00% to 83.81% | 72.45% to 89.36% |
| PPV | 60.27% | 57.97% | 63.42% |
| 95% CI | 50.07% to 69.65% | 48.23% to 67.12% | 51.78% to 73.68% |
| NPV | 89.84% | 89.57% | 87.53% |
| 95% CI | 83.01% to 94.12% | 82.57% to 93.96% | 81.09% to 91.99% |

NPV: Negative predictive value; PPV: positive predictive value
MAXCC=6: disease activity indicated by 6 or more particles in any cross sectional scan; MINCC=3: activity indicated by no fewer than 3 particles in any cross sectional scan; MEDCC=5: activity indicated by median of 5 or more particles per cross sectional scan

### AS-OCT correlation with clinical grading of severity

There was strong correlation between the median cell count per eye, MEDCC, and clinical assessment (Figure 4). This strong correlation was also seen across the other measurands (supplementary figure SF5). However, there was evidence of associations between patient variables and counts, complicating the relationship between slit lamp clinical grading and AS-OCT measurands. Specifically, older individuals, and eyes with heavy pigment loads exhibited higher particle counts when compared to younger or pigment free eyes with a similar clinical SUN grade (table 3). There was no such independent association between particle counts and lens status, ethnicity or sex.

**Figure 4.**
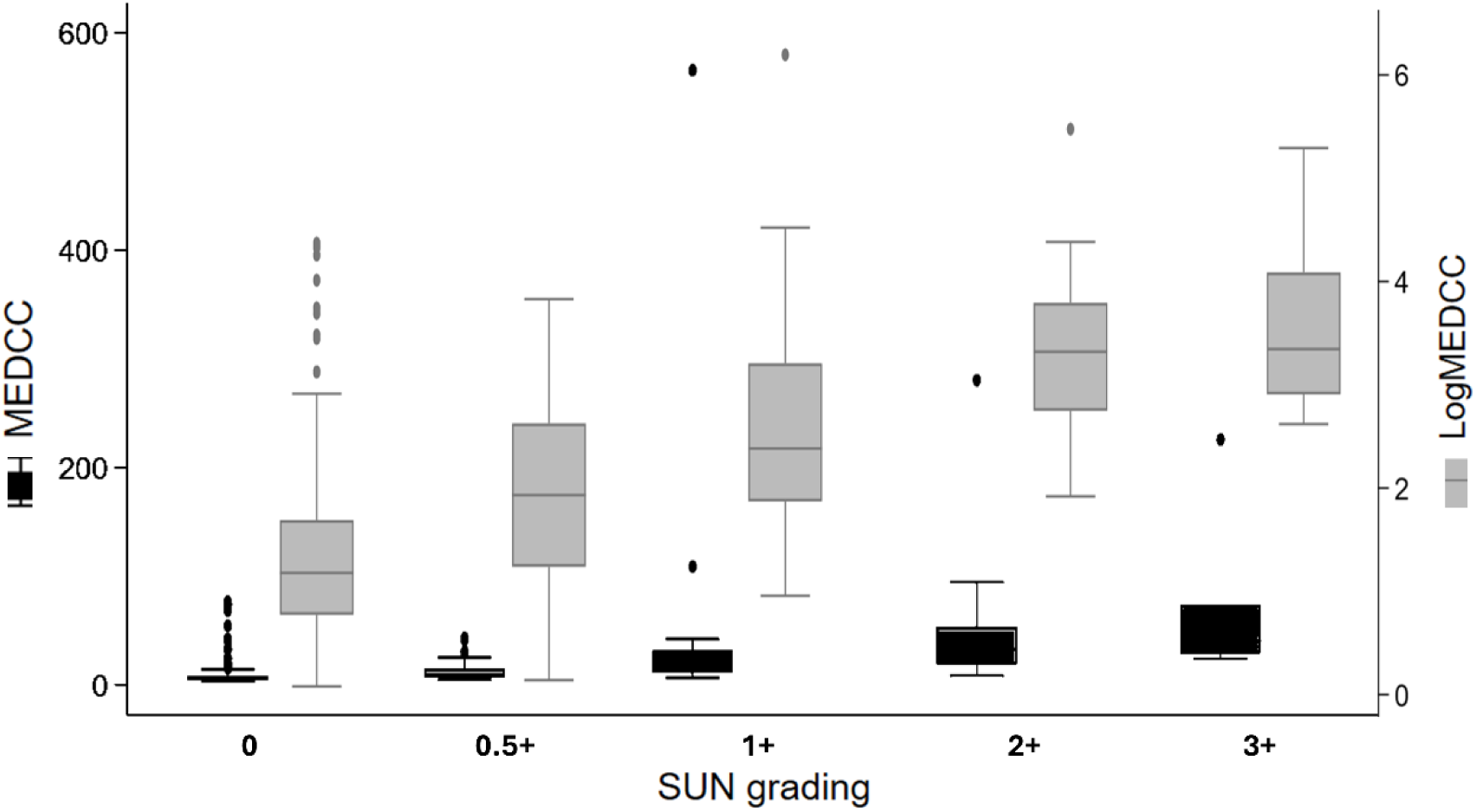
Median cell count per cross sectional image per eye across the different clinical grades. Cell counts across different clinical grades of inflammation shown using actual counts and logarithmic converted units

**Table 3.**
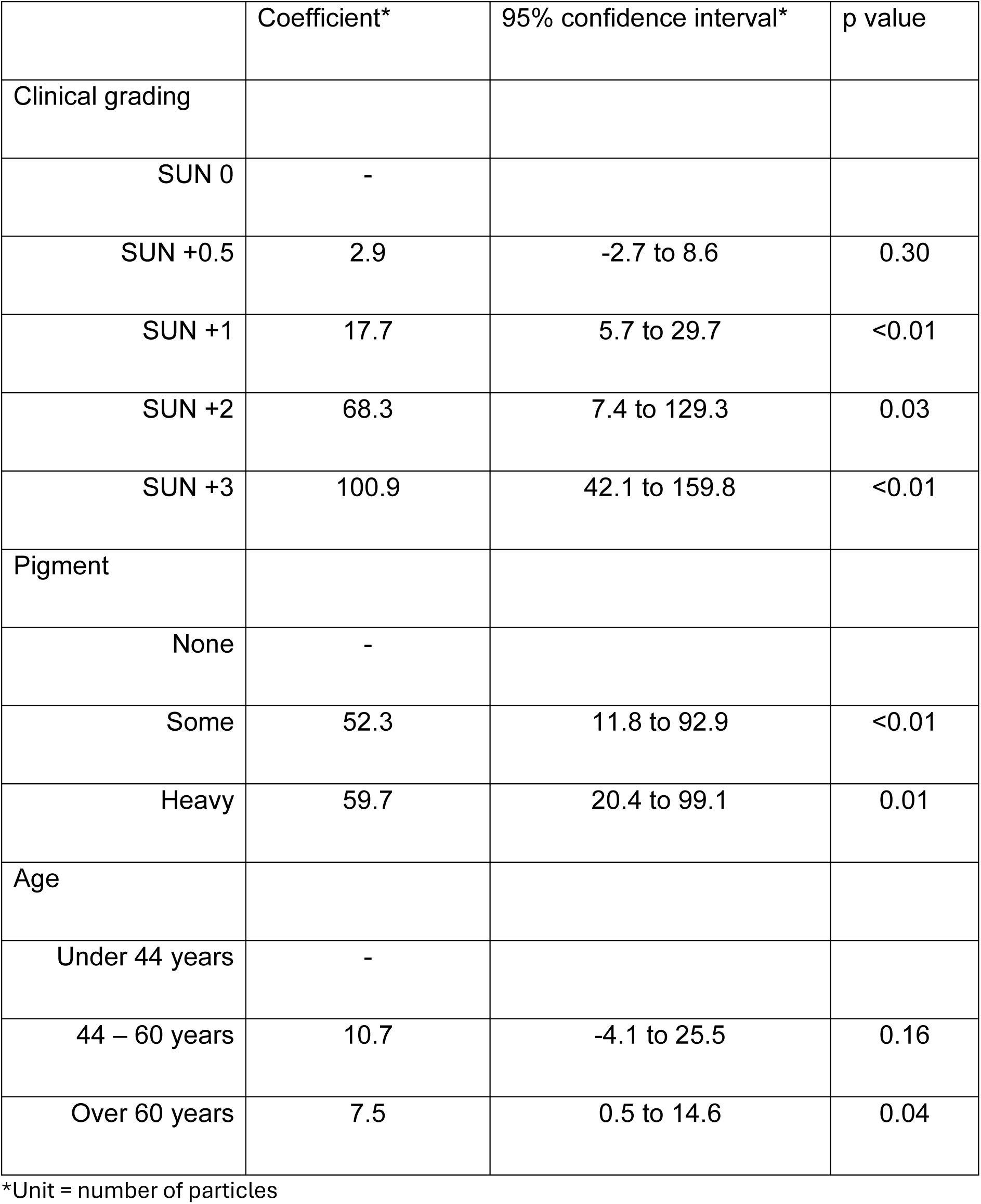
Clinical characteristics associated with MEDCC (median count)

|  | Coefficient* | 95% confidence interval* | p value |
| --- | --- | --- | --- |
| Clinical grading |  |  |  |
| SUN 0 | - |  |  |
| SUN +0.5 | 2.9 | -2.7 to 8.6 | 0.30 |
| SUN +1 | 17.7 | 5.7 to 29.7 | <0.01 |
| SUN +2 | 68.3 | 7.4 to 129.3 | 0.03 |
| SUN +3 | 100.9 | 42.1 to 159.8 | <0.01 |
| Pigment |  |  |  |
| None | - |  |  |
| Some | 52.3 | 11.8 to 92.9 | <0.01 |
| Heavy | 59.7 | 20.4 to 99.1 | 0.01 |
| Age |  |  |  |
| Under 44 years | - |  |  |
| 44 – 60 years | 10.7 | -4.1 to 25.5 | 0.16 |
| Over 60 years | 7.5 | 0.5 to 14.6 | 0.04 |

\*Unit = number of particles

## Discussion

From this cross-sectional analytic and clinical validation study, we report that AS-OCT when using human-in-the-loop automated cell counting with the MCD pipeline, offers a precise, repeatable and clinically accurate method for assessing anterior chamber inflammation in adult patients with uveitis. Automated analyses of images enabled assessment of an otherwise unfeasibly large volume of images and interrogation of various hypotheses of interest. We confirm robust test-retest repeatability, with reproducible measurements across sequential scans, a necessary quality for measures of disease activity. The pipeline was able to differentiate clinical activity from inactivity with high accuracy, and the outputs exhibited strong correlation with SUN grading. Pupil dilation did not significantly impact the precision and variability of AS-OCT measurements of anterior chamber inflammation in our study. There is also stability of performance across populations which differ by lens status, however, AS-OCT derived measurands are typically higher in cell count for older eyes or eyes which had anterior chamber pigments, suggesting that thresholds denoting disease inactivity may need to differ for these populations.

Previous investigators have focused on the ability of AS-OCT to detect or grade disease.^10,16–19^ However, the clinical utility of a quantitative imaging biomarker lies not only in the ability to quantify health or disease, but also in its ability to quantify a meaningful change in that state. Meaningful change is a challenging concept to define for a chronic, relapsing remitting disease such as uveitis. The first challenge to address biomarker utility for disease monitoring is building the evidence base around the limits of precision for that marker.^13^ Sequential testing within this disease cohort suggests a threefold change in particle count is suggestive of a significant, or ‘true’ change in anterior chamber particle load. Our novel findings are a valuable foundation for future longitudinal studies evaluating AS-OCT derived markers of ocular inflammation.

The strong and consistent performance (with regards to reliability and clinical assessment) of different measurands such as minimal cell counts and total cell counts suggests that this technology will perform well across different imaging platforms and different acquisition and analysis protocols. For analyses of swept source OCT images in adults with uveitis, analysis using the ‘least active’ and ‘most active’ cross sectional images may be most useful for ruling out, and ruling in active disease respectively. The performance of the identified measurand thresholds in detecting disease activity amongst those at risk will next need to be tested in prospective studies.

Investigators have postulated an association between AS-OCT hyperreflective particle counts and mydriasis, with particulate matter released into the anterior chamber due to lens-iris friction as the pupil dilates,^20^ despite the apparent stability of other clinical assessments of inflammation severity with dilation.^21^ Whilst we identified consistently higher post dilation cell counts across the different measurands, this difference did not reach statistical significance across the 281 eyes within our study sample. It is noticeable that the prior study recruited older, healthy participants, with a mean age of 73 years (+/- 11 years), versus our younger cohort (median age 48 years, mean 46.3 +/- 13 years). Older, phakic eyes may be more at risk of iris-lens friction due to age related changes in lens morphology,^22^ highlighting the importance of interrogating the performance of novel quantitative techniques across diverse clinical populations.

The impact of anterior chamber pigment load on particle quantification has been previously described.^11^ ‘False positive’ cases, where eyes are thought to be clinically active on AS-OCT due to an anterior chamber containing copious pigment will be important to capture for refinement of this technology. Crucially, ‘false positives’, where hyperreflective are indicative of pigment, or red blood cells, will still be eyes worthy of attention, being cases where identification of acellular material or non-inflammatory cells in the anterior chamber holds clinical utility. This would include eyes which have recently undergone intraocular surgery,^23^ eyes with pigment dispersion disorders,^24^ or anterior chamber blood or metastatic cells.^25,26^ This strengthens the case for implementation of this technology in the first instance as a community-based triage tool. Additionally, the ‘dose-dependent’ relationship between AS-OCT hyper-reflective particles and the most robust metric currently available for grading of inflammation, slit lamp examination by a senior clinician, points to biological plausibility. Longitudinal imaging, which allows assessment of individual relative changes, and associations with clinically meaningful outcomes, will support more confident interpretation.

The positive correlation of age on pre-dilation AS-OCT particle count, which has been identified elsewhere across healthy populations^27^ as well as those with inflammatory eye disease, is less easy to explain. Age related changes in lens morphology,^28^ iris ultrastructure^29^ or in vascular health^30^ could be postulated. Future ophthalmic imaging population epidemiology studies are likely to provide further insight.

Whilst the strength of this study lies in the combination of a large, real world representative sample and the automated analysis pipeline which minimizes operator variability, the work is limited by the absence of the longitudinal data needed to improve the precision of measurement of clinically meaningful change. The use of automated rather than manual analysis has ensured timely completion of image analysis, but the use of such approaches in routine clinical care may prevent wider adoption across lower income settings, or even across higher income settings with challenges in implementing AI based diagnostics. Additionally, while we assessed repeatability, further work is needed to understand AS-OCT reproducibility across multi-center studies and use in clinical trials.^31,32^ Open science principles such as the sharing of the code which underpins this work will support future work across different investigator teams and in turn support wider implementation of this impactful innovation.^33^

In conclusion, AS-OCT offers an objective and reliable method for assessing anterior chamber inflammation in uveitis, with potential applications in both clinical and research settings. Pupil dilation did not appear to have a significant impact on particle count, irrespective of patient characteristics. However, patient characteristics did impact on the correlation of counts with clinical assessment: different thresholds for ‘positive’ AS-OCT detection of activity may be needed for older versus younger patients, and AS-OCT quantification may differ for eyes with heavy anterior chamber pigment loads. The findings presented here will support the future direction of this modality, which sits within a broader movement towards more accessible, technology-enabled remote ophthalmic diagnostics and management. AI-assisted diagnostics are increasingly used to analyze ophthalmic images for earlier detection and monitoring of disease.^34,35^ Tools designed to extend screening and diagnostic capability beyond specialized hospital settings continue to emerge.^36,37^ Future work in this area should focus on refining diagnostic thresholds and evaluating utility in longitudinal studies in order to harness the promise of AS-OCT imaging for populations affected by ocular inflammation.

## Data Availability

All code produced for this study can be found in the following GitHub repository: https://github.com/UCL/udetect.

## Acknowledgements

We are grateful to Gerald Arquisola, Tiffany Wilkinson and the Moorfields Retinal Imaging Team (Benzene Decasa, Bridget Serwaah Akoto, Rikki Rockpowell, Andrew Bakr, Nattapon Boonarpha) for their support of the study. We acknowledge the support from UCL Advanced Research Computing for the technical discussions, computing pipelines, and assistance with establishing reproducible workflows for the open-source code and documentation.

## Data sharing statement

The code used for this study can be found in the following GitHub repository: https://github.com/UCL/udetect.

## Supplementary document SD1. Details of study protocol

### Acquisition device

Specific model: Anterion SS OCT Commercial model: 1300-nm light source and scanning speed of 50,000 A-scans/second and axial depth of 14 mm

### Pupil size variability / standardisation

No formal background lighting assessment – dim room, undilated pupil, default platform fixation light / distance, then again post mydriasis

### Subject positioning

Upright

### Type of scan (line, volume, radial, circular, other)

Volume

### Scan parameters

768 A-scans per B-scan, horizontal; 16-mm scan length, 13 B-scans, centred at the pupil; no averaging

## Supplemental document SD2. Optimisation of inference parameters for the images

A limited subset of B-scans with manual cell annotations was used to assess the behaviour of the pre-trained MCD model when applied to this dataset, which differs from the original training data in signal-to-noise characteristics and spatial resolution. Manual annotation was undertaken by one investigator (TR) and validated using independent review of annotations by a senior clinician (ALS). These annotated images were not used to retrain the model instead, they were used to guide the selection of inference parameters, including detection thresholds and post-processing criteria, that maximised consistency between automated predictions and manual cell counts. While this analysis demonstrated a clear monotonic relationship between annotated and predicted cell counts (Supplemental figure SF1), the limited availability of ground truth annotations and the image-level nature of the labels motivated subsequent validation at the eye level using clinical grading

### Inference

For each B-scan, a zero-shot vision foundation model was first applied to segment the anterior chamber region. Within the segmented AC, MCD employs a Minuscule Region Proposal (MiRP) algorithm to identify candidate regions likely to contain cells, enabling the model to focus computational effort on small, cell-sized patches rather than the full image. Each candidate patch is then passed through a Spatial Attention Network, which learns discriminative local features to distinguish true inflammatory cells from background speckle noise and imaging artefacts.

**Supplemental figure SF1.**
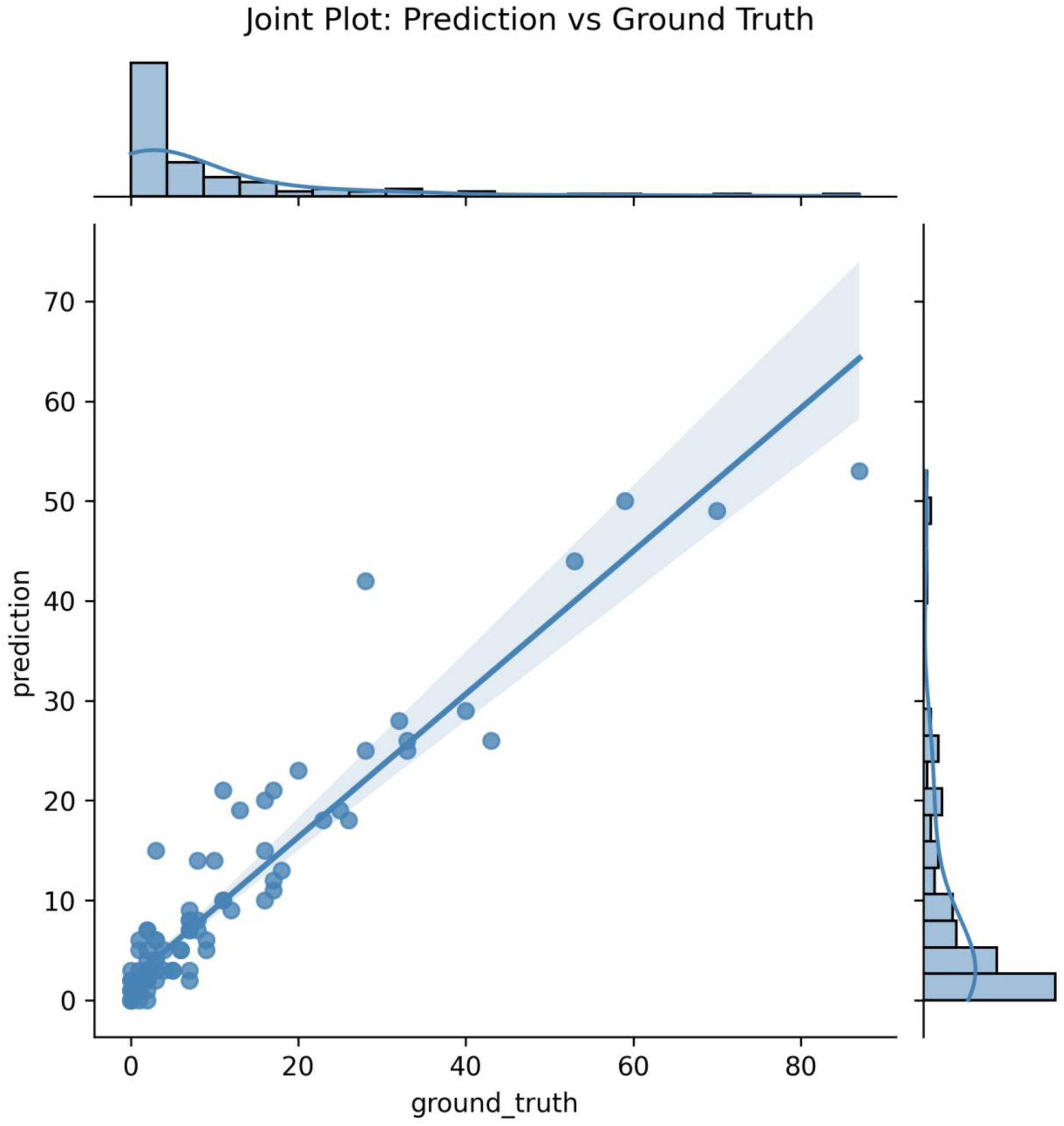
Relationship between manually annotated and predicted cell counts.

**Supplementary Figure SF2.**
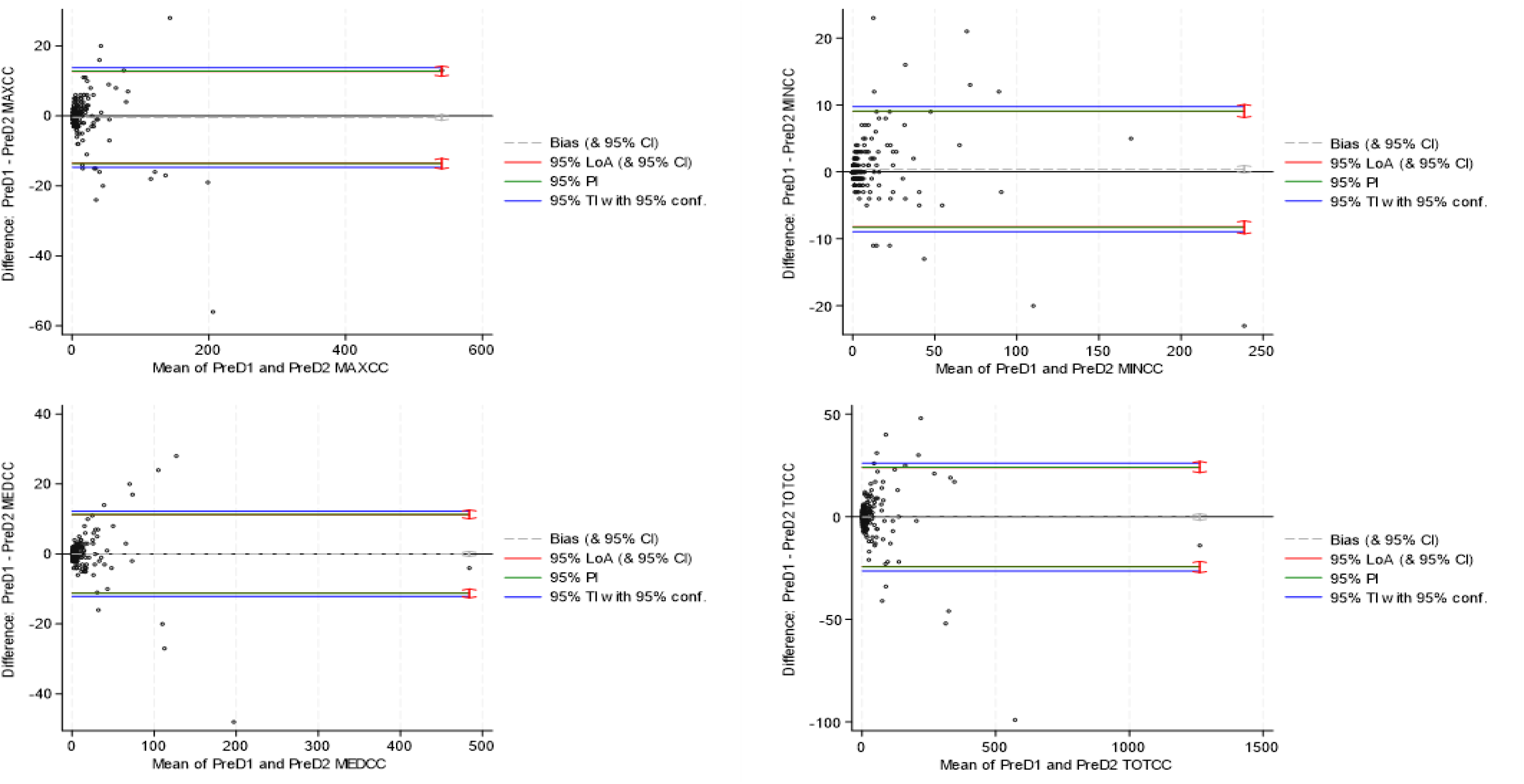
Bland-Altman plots for test-retest variability of measurands, using measurand means and differences. MAXCC: maximum cell count per horizontal cross-sectional scan per volume; MINCC: minimum cell count per cross-sectional scan; MEDCC: median cell count per cross-sectional scan; TOTCC: total cell count per volume; LoA: limits of analysis; PI: performance interval; TI: tolerance interval

**Supplementary Figure SF3.**
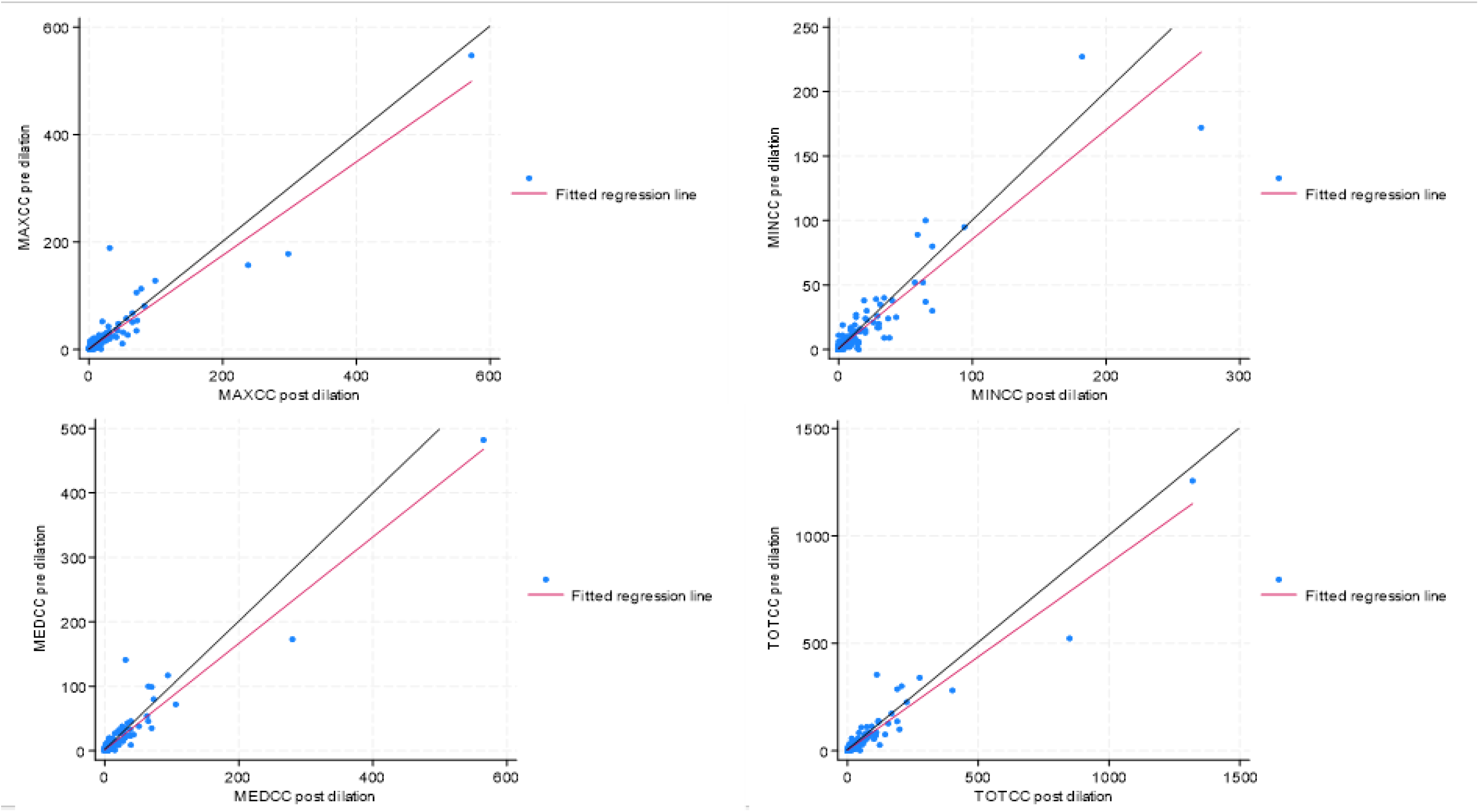
Scatter plots of pre and post dilation variability of measurands. Lines of equivalence shown in black MAXCC: maximum cell count per horizontal cross-sectional scan per volume; MINCC: minimum cell count per cross-sectional scan; MEDCC: median cell count per cross-sectional scan; TOTCC: total cell count per volume

**Supplementary Figure SF4.**
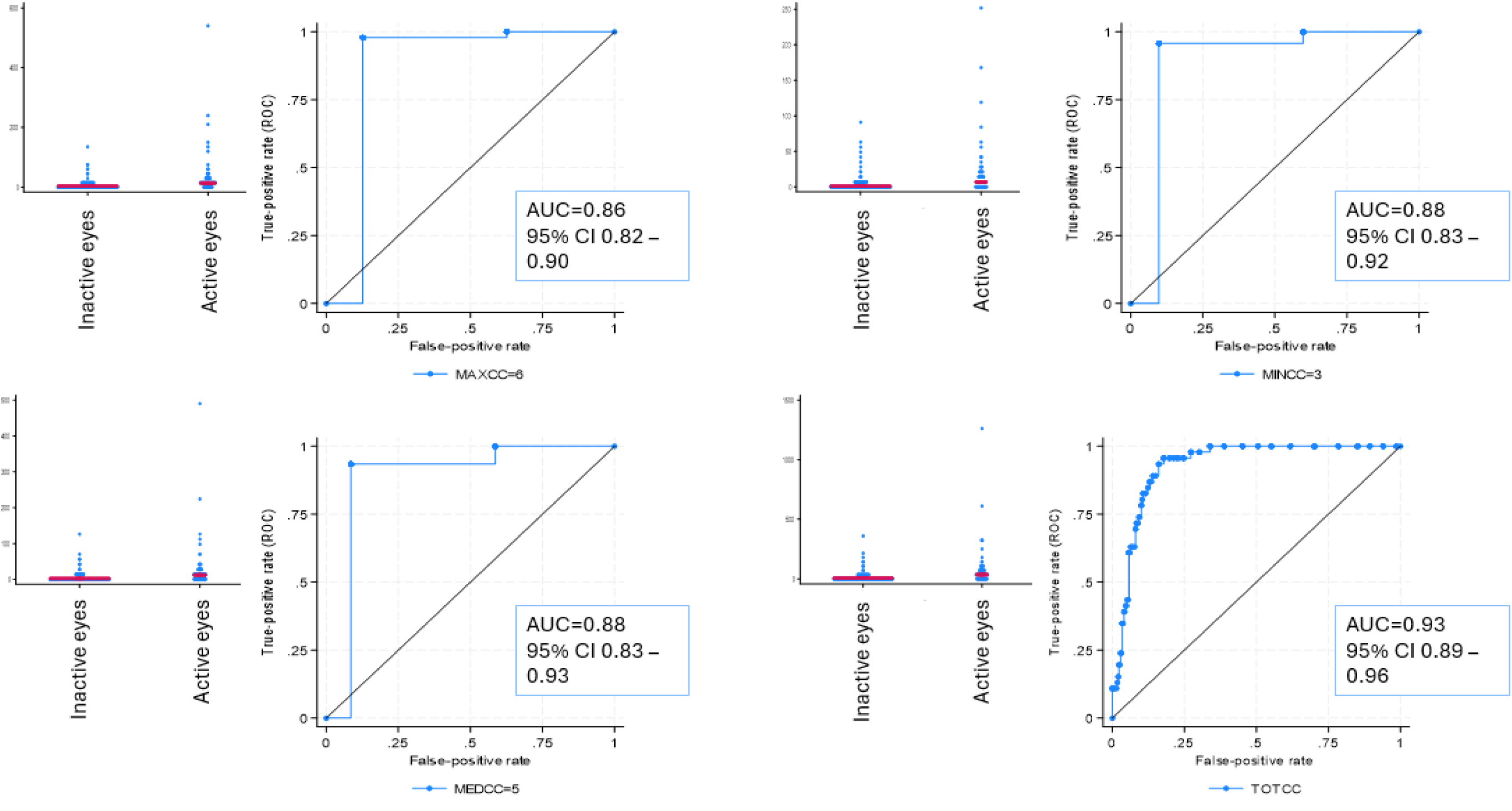
Receiver operated characteristic curves for detection of clinical activity using selected measurand thresholds.

**Supplementary Figure SF5.**
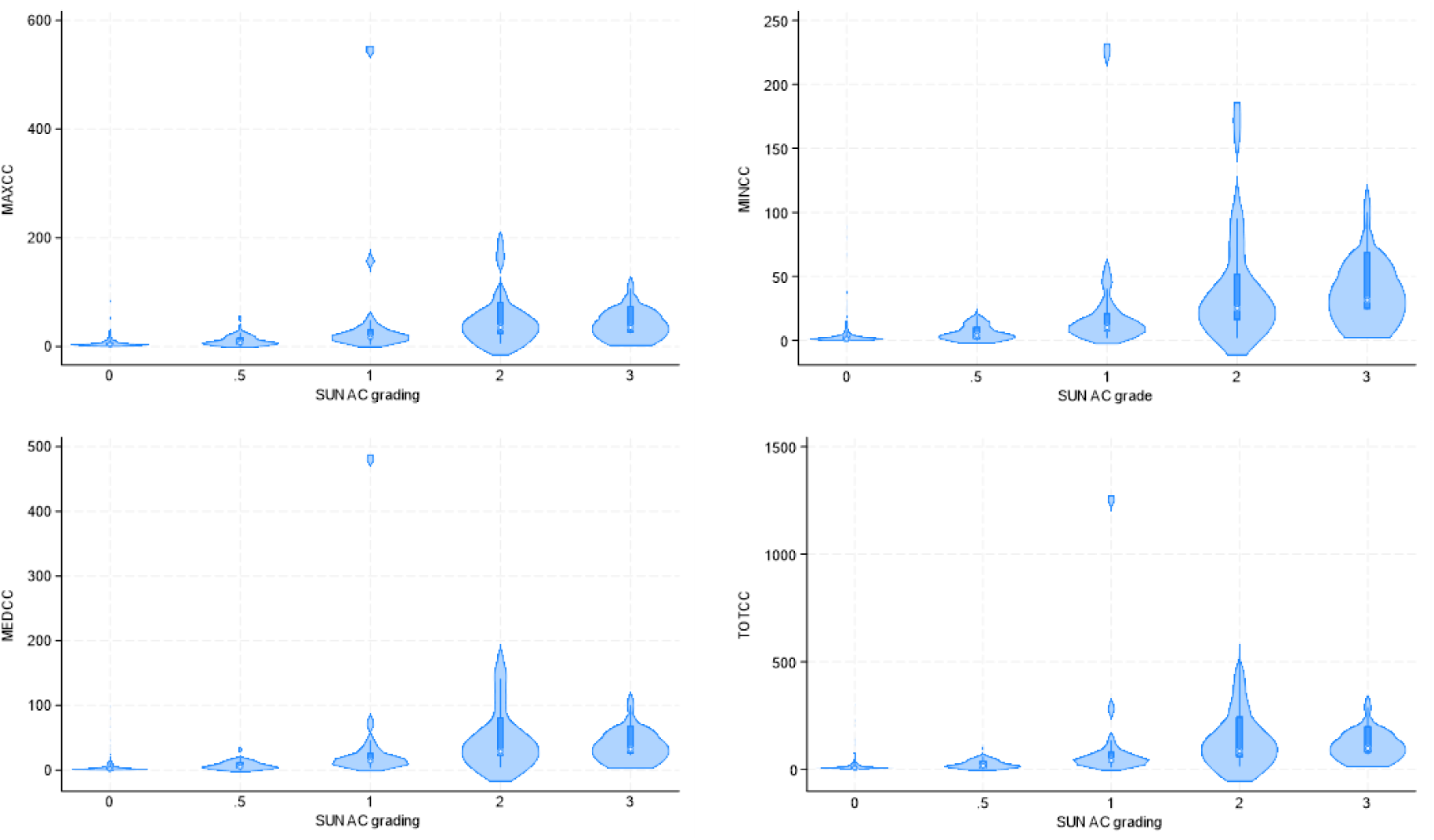
Particle counts quantified using the different measurands across the different clinical grades.

**Supplementary table 1.**
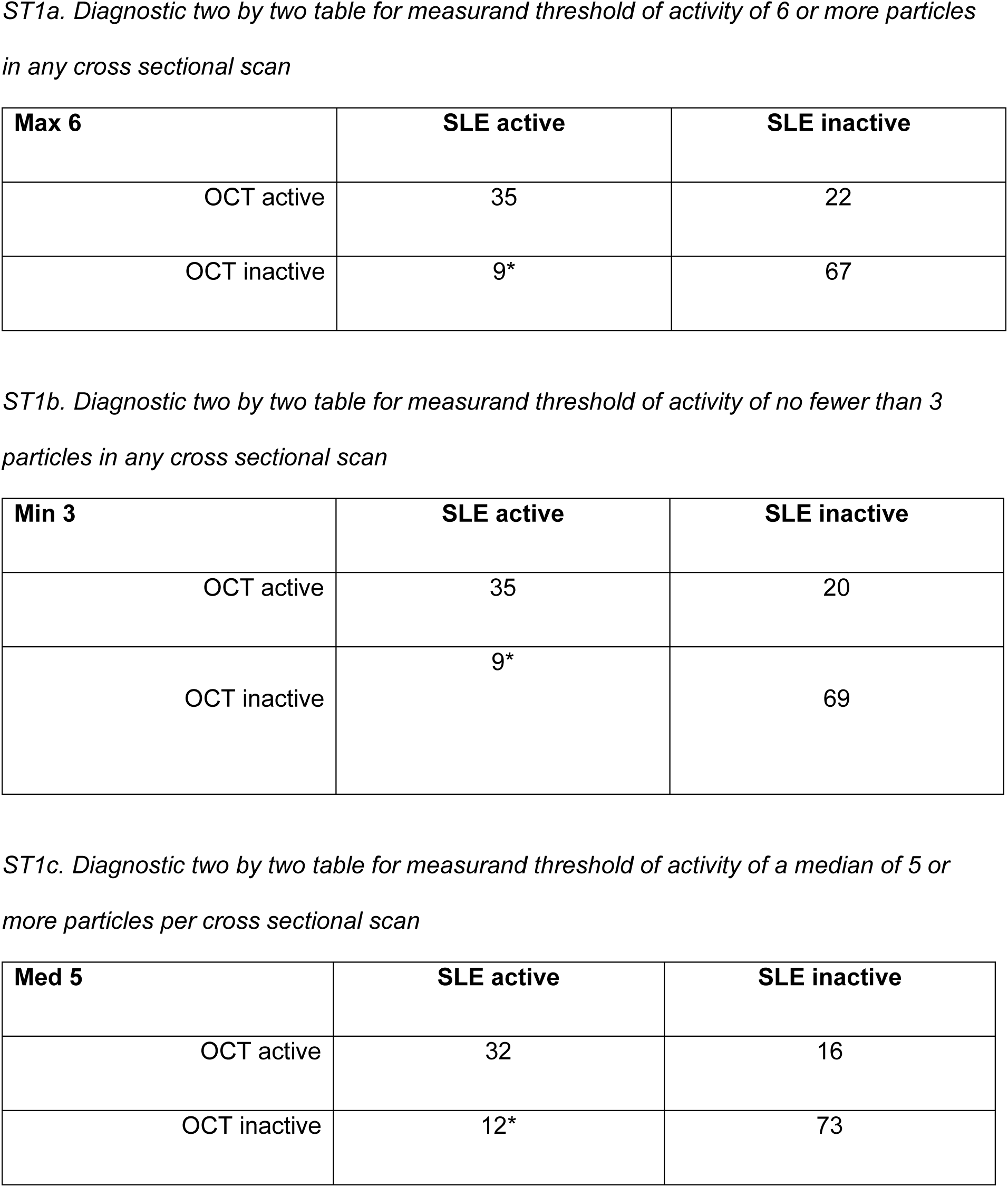
ST1a, ST1b and ST1c: Two by two tables for diagnostic accuracy of selected measurand thresholds.

| **Max 6** | **SLE active** | **SLE inactive** |
| --- | --- | --- |
| OCT active | 35 | 22 |
| OCT inactive | 9* | 67 |

| **Min 3** | **SLE active** | **SLE inactive** |
| --- | --- | --- |
| OCT active | 35 | 20 |
| OCT inactive | 9* | 69 |

| **Med 5** | **SLE active** | **SLE inactive** |
| --- | --- | --- |
| OCT active | 32 | 16 |
| OCT inactive | 12* | 73 |

